# Genetic Insights into Cardiac Conduction Disorders from Genome-Wide Association Studies

**DOI:** 10.1101/2024.11.14.24317316

**Authors:** Bingxun Li, Hongxuan Xu, Lin Wu

## Abstract

**BACKGROUND:** Substantial data support a heritable basis for cardiac conduction disorders (CCDs), but the genetic determinants and molecular mechanisms of these arrhythmias are poorly understood, therefore, we sought to identify genetic loci associated with CCDs.

**METHODS:** We performed meta-analyses of genome-wide association studies to identify genetic loci for atrioventricular block (AVB), left bundle branch block (LBBB), and right bundle branch block (RBBB) from public data from the UK Biobank and FinnGen consortium. We assessed evidence supporting the potential causal effects of candidate genes by analyzing relations between associated variants and cardiac gene expression, performing transcriptome-wide analyses, and ECG-wide phenome-wide associations for each indexed SNP.

**RESULTS:** Analysis comprised over 700,000 individuals for each trait. We identified 10, 4 and 0 significant loci for AVB (*PLEKHA3, TTN, FNDC3B, SENP2, SCN10A, RRH, PPARGC1A, PKD2L2, NKX2-5* and *TBX20*), LBBB (*PPARGC1A, HAND1, TBX5*, and *ADAMTS5*) and RBBB, respectively. Transcriptome-wide association analysis supported an association between reduced predicted cardiac expression of *SCN10A* and AVB. Phenome-wide associations identified traits with both cardiovascular and non-cardiovascular traits with indexed SNPs.

**CONCLUSIONS:** Our analysis highlight gene regions associated with channel function, cardiac development, sarcomere function and energy modulation as important potential effectors of CCDs susceptibility.

---

The cardiac conduction system is crucial for propagating electrical signals that coordinate cardiac contractions to maintain effective blood circulation. Disturbances in this system, termed cardiac conduction disorders (CCDs), can disrupt heart depolarization and even result in bradycardia. Common manifestations of CCDs include sick sinus syndrome (SSS), atrioventricular block (AVB), left bundle branch block (LBBB), and right bundle branch block (RBBB). While significant progress has been made in understanding the molecular mechanisms driving CCDs, current treatments for CCDs is limited and primarily rely on implantable devices such as pacemakers^1^. As a result, further research is needed to enhance our understanding and facilitate the development of pharmacological or genetic therapies^2^.

Although some CCDs can arise secondary to conditions such as ischemia, toxins, or infections^1^, genetic factors also play a significant role in their development, with studies indicating that CCDs and risk for pacemaker insertion cluster within families^3,4^. Some researchers also suggested to include genetic testing as part of the diagnostic workup in AVB patients^5^. In 2021, Thorolfsdottir et al. conducted a genome-wide association study (GWAS) that identified several novel genetic variants associated with SSS, underscoring a strong genetic basis^6^. However, GWAS studies on other major CCDs such as AVB, LBBB, or RBBB, are not yet fully explored. To address this gap, we performed a meta-analysis of GWAS data on AVB, LBBB, and RBBB to gain further insight into the pathogenesis of these disorders.

## Methods

### Study samples and meta-analysis

The study was reported according to STrengthening the REporting of Genetic Association Studies (STREGA) guideline^7^. **Figure 1** depicts the study design. We included individuals with the phenotypes of interest from UK Biobank^8^ and the FinnGen consortium release 11^9^. Details of each cohort as well as download link of GWAS summary statistics are provided in Supplemental Table1. Because those GWAS data are from two independent cohorts, there was no risk sample overlap. We then conducted meta-analysis of included GWAS analyses using a fixed-effects inverse variance weighted approach implemented in METAL^10^, utilizing the default method that incorporates p-values and the direction of effect, weighted by sample size. To address potential population stratification bias, we applied genomic control to all input files. SNPs with allele frequency < 0.5% were excluded from our analysis^11^.

**Figure 1.**
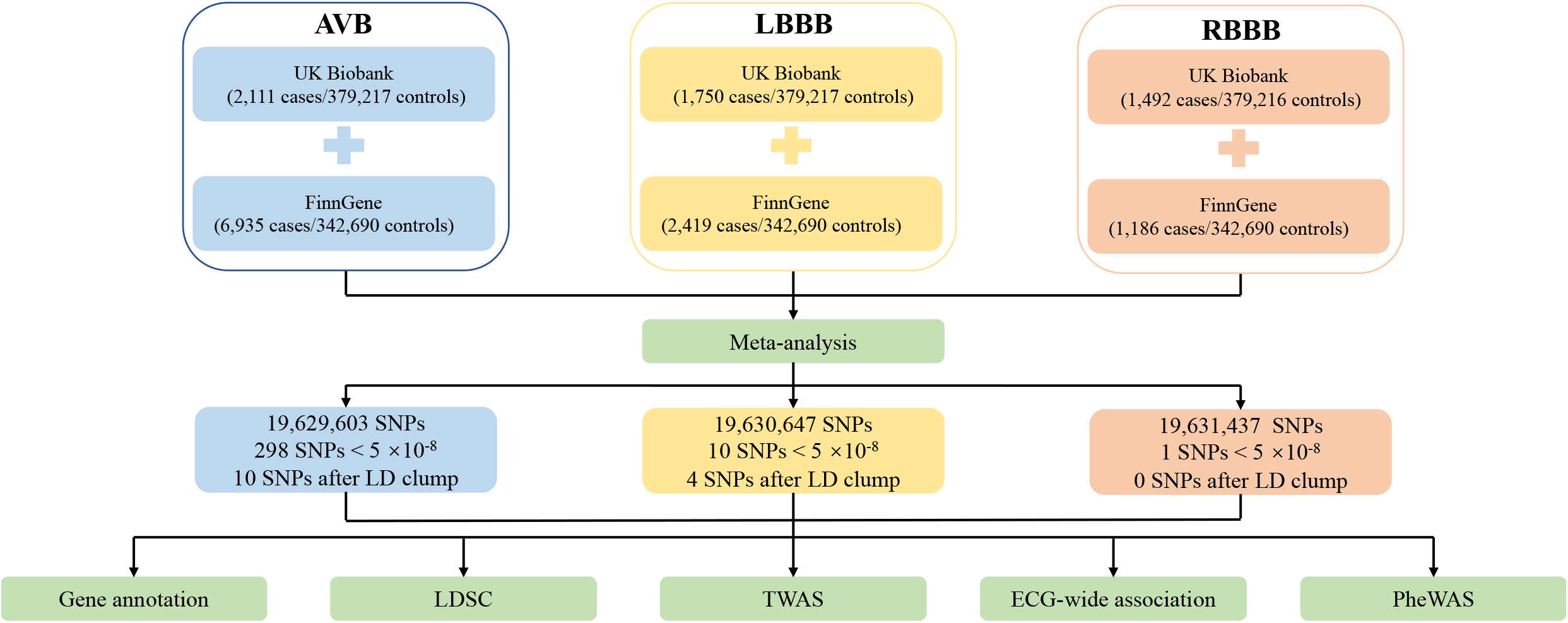
Schematic diagram. AVB, atrioventricular block; LBBB, left bundle branch block; RBBB, bundle branch block; SNP, Single-Nucleotide Polymorphism; LD, linkage disequilibrium; LDSC, LD score regression; PheWAS, phenome-wide association study; TWAS, transcriptome-wide association study

Because the output file of METAL doesn’t include BETA and SE (standard error), which are essential for further analysis, we calculated the above parameters from the following formulations^12^:

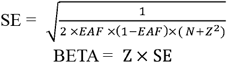

where EAF is effect allele frequency, N is sample size, and Z is Z score.

### Identification of independent locus and novel genes

We used the 1000 Genomes Project phase 3 European (EUR) sample for linkage disequilibrium (LD) estimation via the plinkbinr (version 0.0.0.9) and ieugwasr (version 1.0.2) packages in R (version 4.1.1). Independent locus were defined using a strict standard (clump window = 1Mb, r^2^□=□0.001 and p-value < 5 × 10^−8^). Novel locus were defined as variants more than 500 kb from previously reported ones. We identified the gene closest to the indexed SNP according to Ensembl within a 500 KB region. CADD (Combined Annotation Dependent Depletion) scaled score for each SNP was looked up in the CADD v1.7 website (https://cadd.gs.washington.edu/snv)^13^.

### Transcriptome-wide association study

We applied transcriptome-wide association study (TWAS) to identify significant expression-trait associations using expression imputation from genetic data or summary statistics with FUSION software^14^. This software trains predictive models of the genetic component of a functional or molecular phenotype and predicts and tests that component for association with disease using GWAS summary statistics. FUSION software was used to generate SNP weight sets from BLUP, BSLMM, LASSO, Elastic Net, and top SNPs utilizing genotype and expression data unless BLUP/BSLMM was eliminated owing to sample size or convergence issues. Our goal was to identify associations between the phenotypes of interest and a functional phenotype measured only in reference data; for this analysis, we selected the GTEx version 8 heart left ventricle data^15^. A false discovery rate (FDR) < 0.05 was considered significant. In addition, we conducted colocalization analysis for results with p-value < 0.05 using the coloc package (version 5.1.1)^16^. Colocalization was assessed via posterior probability for hypothesis 4 (PP.H_4_), with PP.H_4_ > 80% indicating significant shared causal variants. We also conduct the joint/conditional tests for further verification.

### LD score regression

We used LD Score regression (LDSC)^17^ to estimate genetic correlations between AVB, LBBB, and RBBB with 9 cardiovascular traits, including Brugada syndrome^18^, heart failure (HF)^19^, coronary heart disease (CAD)^20^, atrial fibrillation (AF)^21^, SSS^6^, QRS duration (from Pan-UKBB GWAS), PR interval^22^, systolic blood pressure (SBP)^23^ and heart rate (HR)^24^ utilizing the ldscr package (version 0.1.0) in R. European LD scores were obtained from the 1000 Genomes Project Phase 3 data for HapMap2 SNPs. A p-value below 0.0019 (0.05/9/3) was considered significant for LDSC. Additionally, we also analyzed the genetic associations of each clumped SNP against those risk factors.

### Phenome-wide Mendelian randomization

To perform Phenome-wide Mendelian randomization (PheWAS) analysis, we queried the GWAS atlas database (https://atlas.ctglab.nl)^25^, which contains 4756 GWAS summary statistics for each indexed SNP. P-values < 7.51□×□10^−7^ (0.05/4756/14) was considered as significant. For those PheWAS results fulfilled the adjusted p-value, we also conducted colocalization analysis for further verification.

### ECG-wide association analyses

Clumped variants of LBBB and AVB GWAS data were looked up in ECGenetics (http://www.ecgenetics.org)^26^ to explore their effect on R-R adjusted three-lead exercise electrocardiogram (ECG) morphology. The GWAS for ECG morphology consists of comprehensive deep phenotyping of 77,190 ECGs in the UK Biobank across the complete cycle of cardiac conduction by plotting 500 association signals of each datapoint as −log_10_(p-values) along the time axis of one heartbeat. This analysis aims to assess the impact of clumped LBBB and AVB variants on ECG morphology, leveraging extensive phenotyping data from the UK Biobank. By visualizing the association signals as −log_10_(p-values), the study seeks to elucidate the relationship between genetic variants and cardiac conduction throughout a complete heartbeat.

## Results

### Meta-analysis results and gene annotation revealed novel genes for AVB and LBBB

We conducted meta-analyses of genome-wide association results for AVB, LBBB, and RBBB using data from the UK Biobank and FinnGen consortium. After quality control, we analyzed 19,629,603 SNPs for AVB, 19,630,647 SNPs for LBBB, and 19,631,437 SNPs for RBBB, respectively. Manhattan plots for each trait are shown in **Figure 2**, while quantile-quantile (QQ) plots and lambda values (**Supplementary Figure 1**) indicated no evidence of inflation. We identified 298 SNPs for AVB, 10 for LBBB, and 1 for RBBB reached GW-significance (i.e., p-value□<□5□×□10^−8^). After LD clumping, 10 SNPs remained for AVB (2 novel SNPs), 4 SNPs for LBBB (2 novel SNPs), and none for RBBB. Details of clumped SNPs are listed in **Table 1**. We identified 10 loci for AVB and 4 loci for LBBB, respectively, in which 2 novel loci for both AVB (rs56065557, rs7433306) and LBBB (rs1472095, rs9636578) were identified.

**Table 1.** Locus reported for AVB and LBBB in the meta-analysis of UK Biobank and FinnGene GWAS datasets. Findings were identified using effects inverse-variance weighted meta-analysis. The chromosomal position is based on GRCh37/hg19 reference. Gene names are ed. Novel locus (i.e., distance > 500kB from previous locus) are shown in bold. CHR, chromosome; POS, base pair location; NEA, fect allele; EA, effect allele; EAF, effect allele frequency; SE, standard error; CADD, Combined Annotation Dependent Depletion.

| SNP | CHR | POS | EA | NEA | EAF | N | BETA | SE | p-value | nearest gene | CADD scaled score |
| --- | --- | --- | --- | --- | --- | --- | --- | --- | --- | --- | --- |
| **AVB** |  |  |  |  |  |  |  |  |  |  |  |
| rs185591978 | 2 | 178488129 | A | G | 0.94 | 349625 | -0.03 | 0.005 | $2.60 \times 10^{-9}$ | *PLEKHA3* | 1.44 |
| rs13031826 | 2 | 178891875 | A | T | 0.88 | 732669 | -0.02 | 0.003 | $1.19 \times 10^{-12}$ | *TTN* | 4.28 |
| rs10936712 | 3 | 172084486 | T | C | 0.69 | 349625 | -0.01 | 0.003 | $1.48 \times 10^{-8}$ | *FNDC3B* | 3.09 |
| **rs56065557** | **3** | **185636428** | **C** | **G** | **0.70** | **732669** | **-0.01** | **0.002** | **$8.35 \times 10^{-9}$** | ***SENP2*** | **1.44** |
| **rs7433306** | **3** | **38729148** | **C** | **G** | **0.57** | **732669** | **0.01** | **0.002** | **$1.45 \times 10^{-10}$** | ***SCN10A*** | **0.29** |
| rs199561133 | 4 | 110760807 | T | G | 0.86 | 349625 | -0.02 | 0.003 | $2.59 \times 10^{-8}$ | *RRH* | 0.57 |
| rs2932971 | 4 | 23817261 | T | C | 0.71 | 732669 | 0.01 | 0.002 | $1.89 \times 10^{-10}$ | *PPARGC1A* | 19.3 |
| rs76734697 | 5 | 137897780 | T | G | 0.81 | 732669 | 0.01 | 0.002 | $2.73 \times 10^{-9}$ | *PKD2L2* | 3.15 |
| rs6556061 | 5 | 173218944 | A | T | 0.72 | 349625 | -0.02 | 0.003 | $2.62 \times 10^{-9}$ | *NKX2-5* | 0.11 |
| rs11764098 | 7 | 35466100 | C | G | 0.97 | 732669 | -0.04 | 0.005 | $2.52 \times 10^{-13}$ | *TBX20* | 18.1 |
| **LBBB** |  |  |  |  |  |  |  |  |  |  |  |
| **rs1472095** | **4** | **23816454** | **T** | **C** | **0.71** | **727785** | **0.010** | **0.002** | **$3.35 \times 10^{-8}$** | ***PPARGC1A*** | **3.29** |
| rs13185595 | 5 | 154492610 | A | G | 0.63 | 727785 | -0.012 | 0.002 | $5.55 \times 10^{-13}$ | *HAND1* | 7.93 |
| rs61931006 | 12 | 114414703 | A | C | 0.90 | 727785 | 0.018 | 0.003 | $4.10 \times 10^{-11}$ | *TBX5* | 2.64 |
| **rs9636578** | **21** | **27538384** | **A** | **G** | **0.52** | **727785** | **-0.001** | **0.002** | **$4.02 \times 10^{-8}$** | ***ADAMTS5*** | **2.94** |

**Figure 2.**
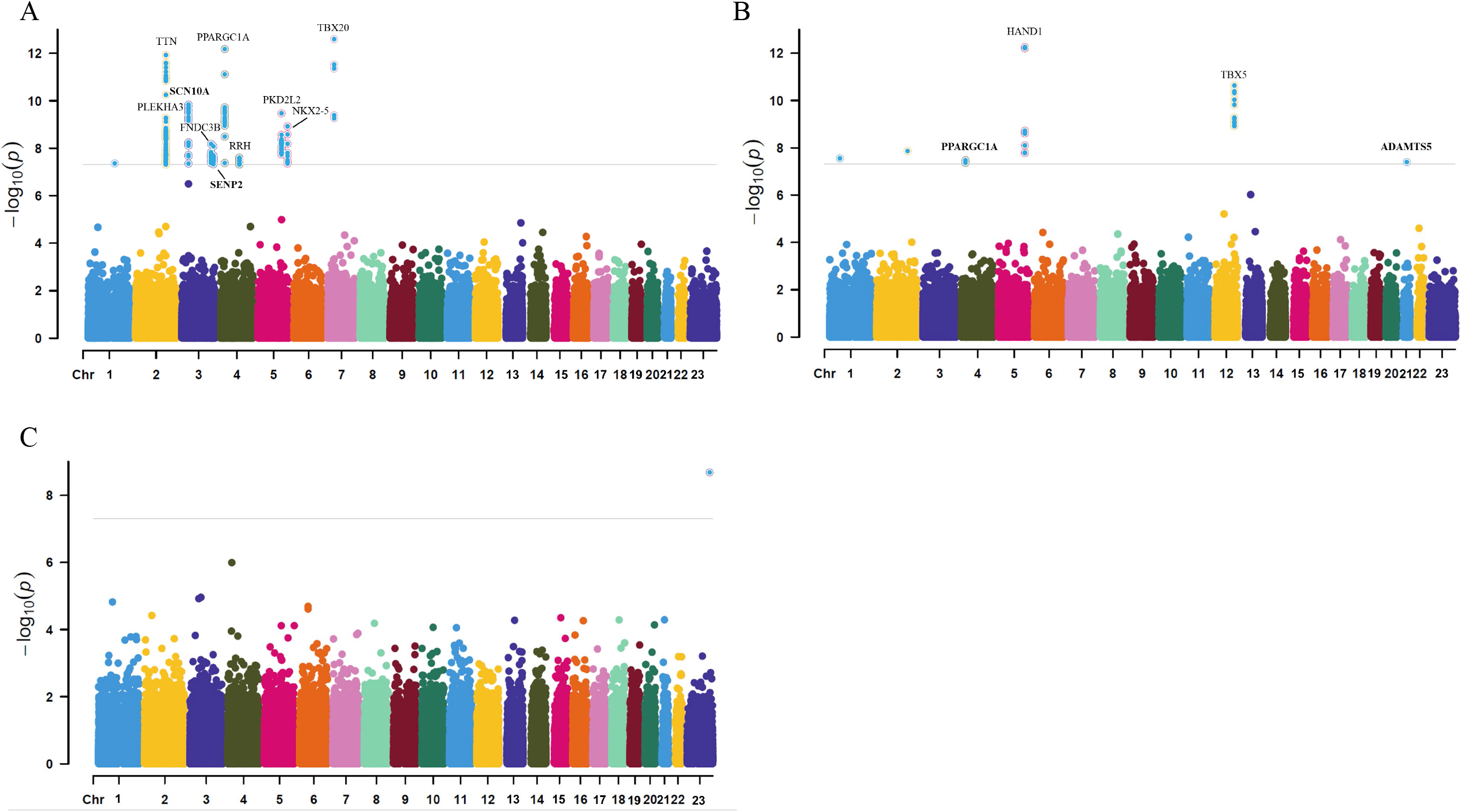
Manhattan plots showing GWAS meta-analysis of. A) AVB; B) LBBB; and C) RBBB. Those plots shows the −log_10_(p-value) of association for each SNP from the GWAS meta-analysis plotted on the y-axis against genomic position on the x-axis. The gray line corresponds to the genome-wide significance threshold (5□×□10^−8^). Novel locus identified genes are highlighted in bold.

We then mapped potential genes according to distances on each chromosome for indexed SNPs. We identified *PLEKHA3, TTN, FNDC3B, SENP2, SCN10A, RRH, PPARGC1A, PKD2L2, NKX2-5, TBX20* as potential genes for AVB indexed SNPs. For LBBB indexed SNPs, the potential genes included *PPARGC1A, HAND1, TBX5*, and *ADAMTS5*.

### TWAS analysis indicated *SCN10A* as potential modulator for AVB

In the single-tissue internal validation using FUSION, of the 5,886 genes included in our genotype data showing significant cis-genetic expression in whole blood according to the GTEx dataset, 10 and 2 genes revealed significant association signals in the TWAS with a FDR p-value less than 0.05 for AVB and LBBB, respectively, as detailed in **Supplementary Table S2** and illustrated in **Supplementary Figure 2**. No significant results were obtained for RBBB GWAS. After colocalization analysis, there were only 4 genes with PP.H4 > 0.80. In subsequent joint/conditional tests, we found that after conditioning on ENSG00000185313.6 (*SCN10A*) in the AVB GWAS data on chromosom 3, in which this locus goes from being genome-wide significant to non-significant after conditioning on the predicted expression of ENSG00000185313.6 (*SCN10A*), as shown in **Supplementary Figure 3**. The other 3 genes didn’t have any locus with strong enough signals to visualize due to weak effects or low statistical power.

### Genetic correlation estimates showed associations between CCDs with cardiovascular traits

Estimates of the genetic correlation between AVB, LBBB, RBBB and 9 risk factors are reported in **Table 2**, in which only results with p-value < 0.0019 are listed. We found that CAD and AF were closely correlated with all 3 CCDs, and HF was correlated with AVB and LBBB. **Supplementary Figure 4** showed genetic associations of clumped locus s against 9 cardiovascular traits. Notably, while AVB and PR interval exhibited a strong correlation, we did not identify a significant correlation between QRS duration and LBBB/RBBB. This is likely due to the fact that PR interval and AVB are both focused on the atrioventricular node, a specific conduction pathway, whereas QRS duration reflects a broader range of electrical processes within the ventricles. These processes may not always align genetically with the more localized conduction abnormalities in LBBB and RBBB. The difference in complexity and specificity of these traits explains why a strong genetic correlation was found in the former pair but not in the latter. Additionally, we found that BrS and RBBB didn’t share as much strong genetic correlation as expected. This might indicate that although initially reported as a RBBB-like ECG change^27^, BrS and RBBB are distinct diseases with different genetic background.

**Table 2.** LDSC analysis results showing genetic correlations between CCDs and related cardiovascular traits. Here only significant results were present. AVB, atrioventricular block; LBBB, left bundle branch block; RBBB, right bundle branch block; AF, atrial fibrillation; CAD, coronary heart disease; HF, heart failure; SSS, sick sinus syndrome; SBP, systolic blood pressure; *r*_g_, genetic correlation; SE, standard error.

| Trait1 | Trait2 | $r_g$ | SE | p-value |
| --- | --- | --- | --- | --- |
| AVB | AF | 0.46 | 0.07 | $6.86 \times 10^{-10}$ |
| | CAD | 0.30 | 0.05 | $1.18 \times 10^{-9}$ |
| | HF | 0.48 | 0.08 | $5.17 \times 10^{-10}$ |
| | SSS | 0.32 | 0.07 | $5.85 \times 10^{-6}$ |
| | PR | 0.76 | 0.17 | $5.41 \times 10^{-6}$ |
| | HR | 0.19 | 0.04 | $1.98 \times 10^{-5}$ |
| LBBB | AF | 0.26 | 0.07 | $1.19 \times 10^{-4}$ |
| | CAD | 0.40 | 0.06 | $7.59 \times 10^{-12}$ |
| | HF | 0.65 | 0.10 | $2.83 \times 10^{-10}$ |
| | SBP | 0.24 | 0.05 | $7.16 \times 10^{-6}$ |
| RBBB | AF | 0.36 | 0.11 | $1.10 \times 10^{-3}$ |
| | CAD | 0.51 | 0.09 | $2.50 \times 10^{-8}$ |

### ECG-wide associations

The influence of indexed SNPs on ECG morphology is shown in **Supplementary Figure 5**. We showed the ECG morphology phenotype plots as well as the heatmaps for the indexed SNPs. The plots showed that the genetic ECG signature of AVB clumped SNPs mainly affected the PR-segment morphology on the ECG, and LBBB indexed SNPs mainly affected the QRS wave morphology.

### PheWAS analysis

The results of PheWAS were present in **Supplementary Table 5**. We found that apart from AF and some common ECG traits such as PR interval, HR, the indexed SNPs in our analysis were also to be highly associated with other non-cardiac traits, such as estimated glomerular filtration rate, impedance measures, body anthropometric traits, etc. Because the GWAS data of ECG traits from Genevieve Wojcik et al^28^ were not from individuals of European ancestry, we excluded those from our results and conducted colocalization analysis. After colocalization analysis, we identified that some traits with high colocalization evidence (PP.H4 > 0.80) with indexed SNPs, and the virtualization of those results were present in **Supplementary Figure 6 - 11**.

### Discussion

In this meta-analysis of GWAS for 3 clinically important CCDs using 2 independent cohorts, we identified 10 and 2 genome-wide disease susceptibility loci for AVB and LBBB, respectively, including 2 novel locus were identified for each GWAS. Our findings primarily highlighted genes involved in ion channel function, sarcomere function, cardiac development and energy modulation, supporting and extending previous findings suggesting an genetic basis of CCDs and underscore the complex genetic architecture underlying these disorders^3-6^. Additionally, they suggest potential pathways for future research and therapeutic targeting.

Most susceptibility locus are associated with genes that affect cardiac ion channels, such as *SCN10A, TBX20* and *TBX5*. Variations in *SCN10A* have been identified to influence cardiac conduction and PR interval^29^. TWAS analysis also suggested *SCN10A* as an important regulator for AVB. The T-box proteins are also essential for cardiac conduction system morphogenesis and activation or repression of key regulatory genes^30^. These genes primarily impact the cardiac conduction system, potentially leading to conduction delays.

We also observed susceptibility locus for both AVB and LBBB that mapped to *PPARGC1A* (also known as *PGC-1*α), an important mitochondrial energy metabolism factor, suggesting that *PPARGC1A* may be an important modulator for cardiac conduction. *PPARGC1A* is involved in the regulation of oxidative phosphorylation and mitochondrial biogenesis, processes critical for maintaining cellular energy homeostasis, and is closely associated with cardiovascular diseases^31^. Disruption of mitochondrial function can lead to altered ion channel expression and disrupted Ca^2^□ homeostasis, which in turn increases susceptibility to arrhythmias in both atrial^32^ and ventricular tissues^33^ in mice with *PPARGC1A* deficiency.

Our results further implicate the cardiac sarcomere for AVB and LBBB. We identified a susceptibility locus for AVB at *PLEKHA3*, located just downstream of *TTN*. TTN encodes the large cardiac structural protein titin, which is essential for the assembly and function of vertebrate striated muscles. By providing connections at the level of individual microfilaments, titin helps maintain the delicate balance of forces between the two halves of the sarcomere. Variants in TTN are among the most common inheritable risk factors for dilated cardiomyopathy^34^ and have been associated with early-onset AF^35^. A recent research also identified TTN as an important susceptibility locus paroxysmal supraventricular arrhythmia related to accessory pathways^36^.

We also found some genes associated with cardiac development. The Small Ubiquitin-like Modifier (SUMO) modification, implicated in various cellular processes including protein trafficking, transcriptional regulation, protein stability, cell death, and survival, is important to maintain physiological function. SUMOylation of several ion channels such as SCN5A to affect conduction^37^. We found SENP2, a key member of the SENPs family that regulates SUMOylation, in AVB susceptibility locus. The downstream target of SENP2 including NKX2-5^38^, another identified gene in AVB GWAS data, which functions in heart formation and development. Mutations in this gene cause atrial septal defect with atrioventricular conduction defect, and also tetralogy of Fallot, which are both heart malformation diseases^39^. Another gene with potential influence on cardiac development is HAND1 as one of LBBB susceptibility locus. HAND1 is a key transcription factor involved in early cardiogenesis. It is one of two closely related family members known as the HAND proteins, which are asymmetrically expressed in the developing ventricular chambers and play an essential role in cardiac morphogenesis. Acting in a complementary manner, these proteins are crucial for the formation of the right ventricle and the aortic arch arteries, implicating them as significant mediators of congenital heart disease^40^.

We also observed some genes, such as RRH, FNDC3B, and ADAMTS5, that have not been extensively studied in the context of cardiovascular function. Additionally, our PheWAS analysis revealed that some indexed SNPs were closely associated with non-cardiac traits, including estimated glomerular filtration rate (eGFR). These findings suggest a broader role for these genes and SNPs beyond traditional cardiovascular implications, highlighting the need for further investigation into their functions.

### Limitations

We recognize several limitations in our analysis. Firstly, the genetic data primarily derives from individuals of European ancestry, which may restrict the applicability of our findings to other ethnic groups. Secondly, despite a total sample size exceeding 700,000, we did not retain SNPs in the RBBB GWAS data after clumping. Further studies are necessary to address this gap.

## Conclusion

In this meta-analysis of GWAS including 2 independent cohorts and over 700,000 European individuals, we identified 10 and 2 previously reported genome-wide associations with AVB and LBBB, respectively, and 2 novel associations for each GWAS. Our findings primarily implicate genes involved in ion channel function, cardiac development, sarcomere function and energy modulation as important potential effectors of risk for AVB and LBBB.

## Data Availability

All data produced in the present study are available upon reasonable request to the authors

## Data availability

All the data used were acquired from publicly available dataset according to the PubMed ID or links provided in **Stable 1**. No restricted dataset was used in this study. The summary statistics didn’t contain any personal information. For code supporting this manuscript, please refer to https://github.com/bingxunli/block.

## Authors’ contributions

BX Li and HX Xu (first author): Conceptualization, Methodology, Software, Investigation, Formal Analysis, Writing and Original Draft; Y Chen and YY Lin: Data Curation, Writing and Original Draft; L Wu (Corresponding Author): Writing, Review and Editing, Funding acquisition. All authors have reviewed the manuscript and approved its submission in current version.

## Ethics approval

Not applicable

## Consent to participate

Not applicable

## Consent for publication

Not applicable

## Acknowledgments

We would like to extend our deepest appreciation to the providers of open data and all the individuals who participated in the studies.

