## Supplementary material for "Genetic Insights into Cardiac Conduction Disorders from Genome-Wide Association Studies": S Figures

**Supplementary Figures** for "Genetic Insights into Cardiac Conduction Disorders from Genome-Wide Association Studies"

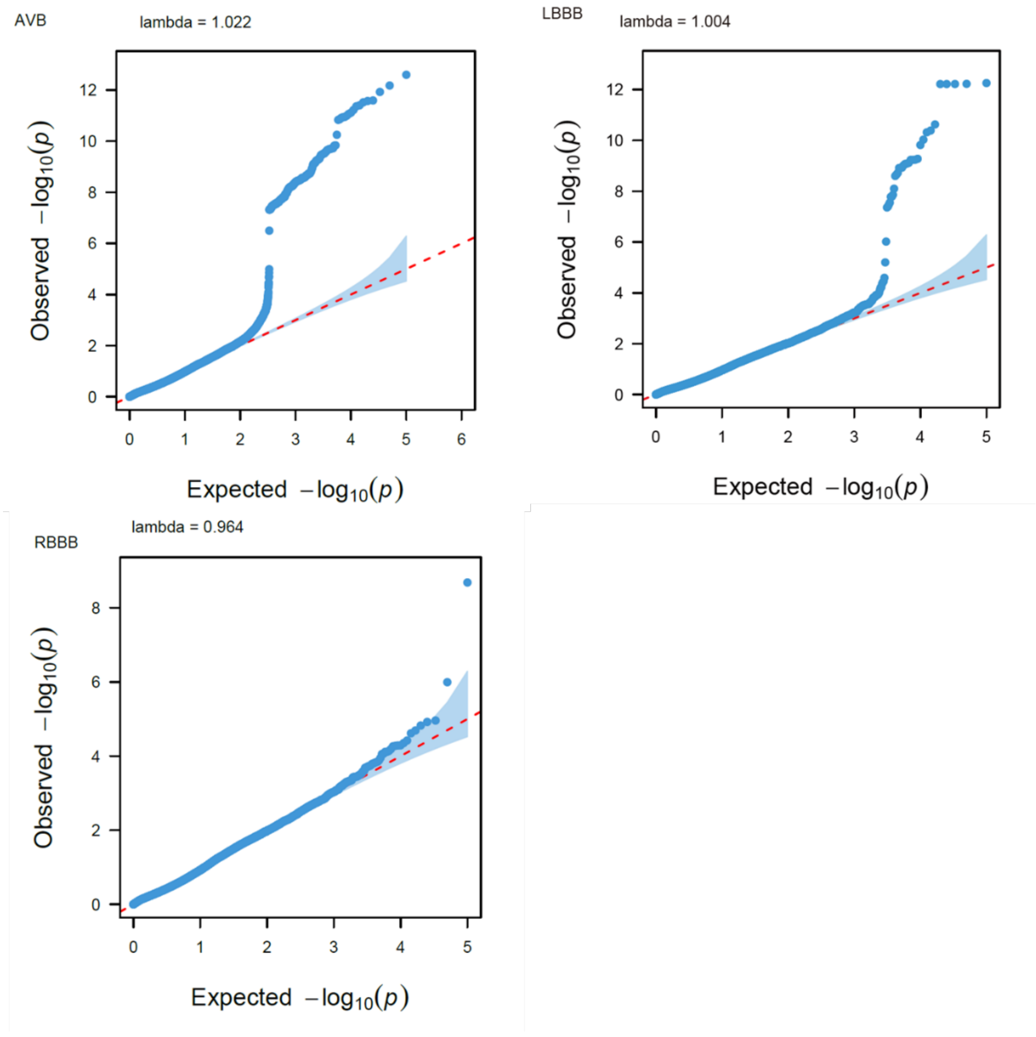


**Supplementary Figure 1**. **Quantile-quantile (Q-Q) plot of CCDs GWAS.**
The Q-Q plot illustrates the number and magnitude of observed associations between genotyped SNPs and CCDs, compared to the expected association statistics under the null hypothesis of no association. The red dashed line represents the identity line. Observed association statistics are plotted on the y-axis, while expected association statistics are on the x-axis, both transformed to -$\log_{10}$(p-value). The lambda value across all Q-Q plots are close to 1, indicating no risk of inflation in the test statistics.

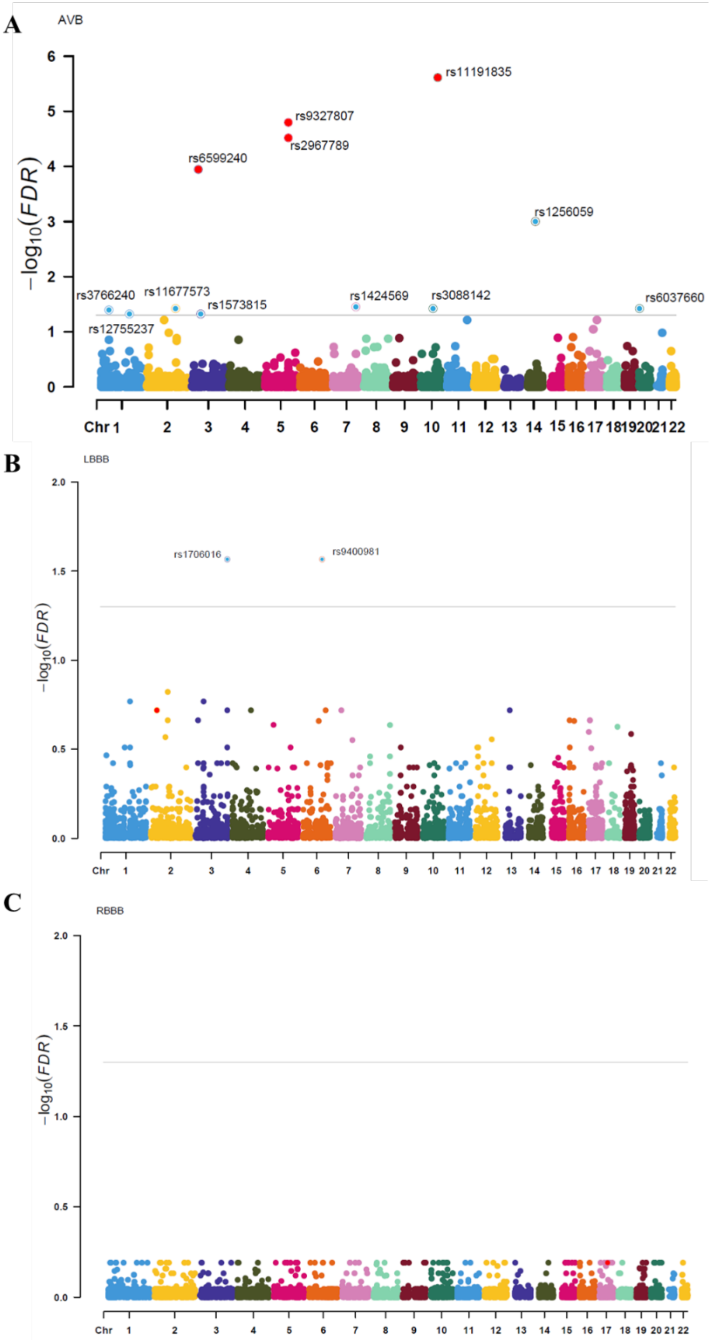


**Supplementary Figure 2. Manhattan plots of TWAS analysis results**

This figure presents Manhattan plots for TWAS analysis of: A) AVB; B) LBBB; and C) RBBB, utilizing FUSION software and GTEx v8 left ventricle heart data. The plots display the -log10(FDR) of associations for each SNP from the GWAS meta-analysis on the y-axis, against genomic position on the x-axis. The gray line indicates the FDR threshold (0.05). SNPs with FDR p-values < 0.05 are shown, with colocalization showing $PP.H_4 > 0.80$ highlighted in red.

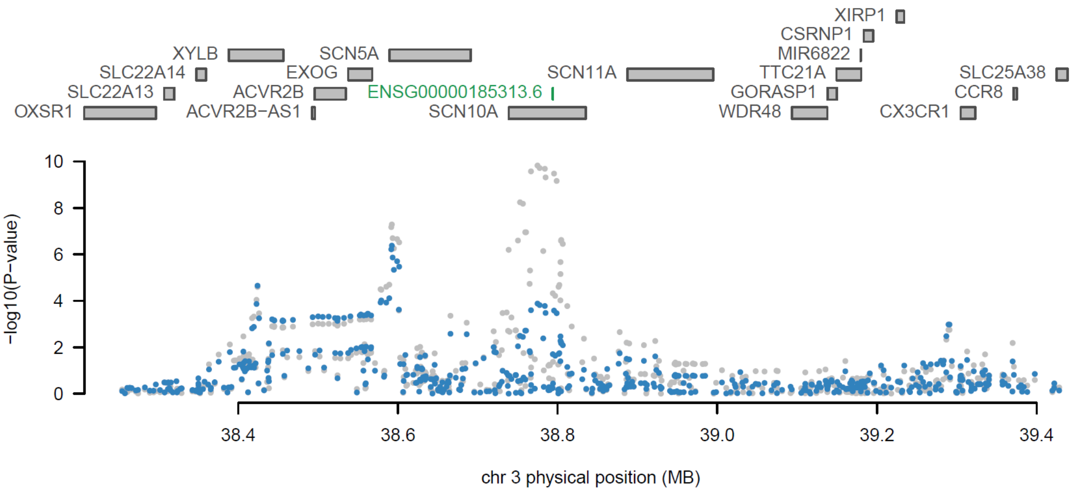


**Supplementary Figure 3. TWAS joint/conditional plots of AVB GWAS data.**
In the top panel, ENSG00000185313.6 is highlighted in green, showing evidence of joint significance with AVB. The bottom panel displays a Manhattan plot of AVB GWAS data before (gray) and after (blue) conditioning on ENSG00000185313.6 (*SCN10A*), in which this locus goes from being genome-wide significant to non-significant after conditioning on the predicted expression of ENSG00000185313.6 (*SCN10A*).

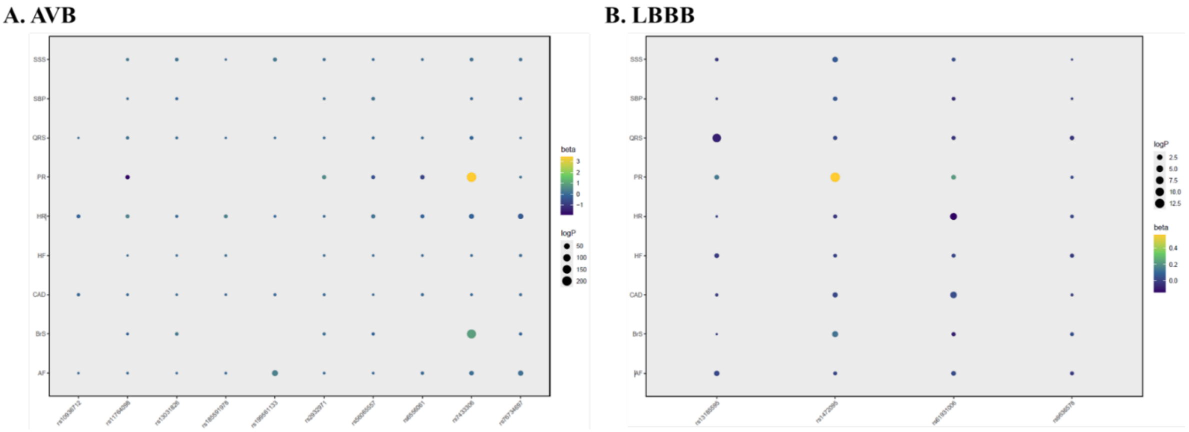


**Supplementary Figure 4. Genetic associations of clumped locis against 9 cardiovascular traits.**
The bubbles represent genetic associations between clumped loci (x-axis) and cardiovascular traits (y-axis). The color of each bubble indicates the beta coefficient of the association: blue represents negative coefficients, while yellow indicates positive coefficients. The size of the bubbles corresponds to the negative logarithm of the association p-value, with larger bubbles indicating lower p-values.

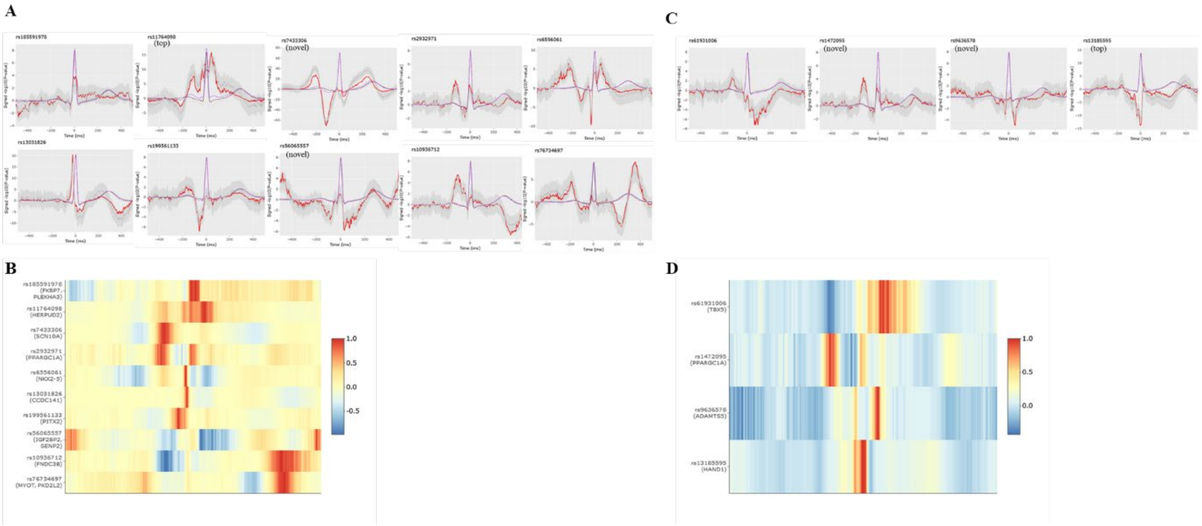

**Supplementary Figure 5. ECG-wide association results of indexed SNPs in the ECGenetics website.**

In the upper panles of Figure A (AVB) and B (LBBB), the left y axis depicts the micro voltage scale, the y axis on the right indicates the signed $-\log_{10}$(p-values), and the x axis is time in milliseconds (ms) from the R-R in the case of the R-R-adjusted ECG morphology phenotype. The blue line is the average ECG amplitude of the full cohort (with 95% confidential interval) and the red line is the p-value for association with each datapoint of the ECG morphology phenotype (n = 500 time points) on a $\log_{10}$ scale, signed to show direction of association. Top refers to top SNP with the smallest SNP, and novel refers to novel SNP with distance > 500Kb from previous GWAS-clumped SNPs. The lower panles of Figure A and B showed heatmaps of the genetic ECG signatures of indexed SNPs of AVB and LBBB, respectively. We normalized the genetic ECG signatures of genetic variants in order to compare the effects across loci. Effects were orientated to the most positively associated allele across all time points and colored in red on the heatmap; a blue color indicates a negative effect while yellow indicates no effect.

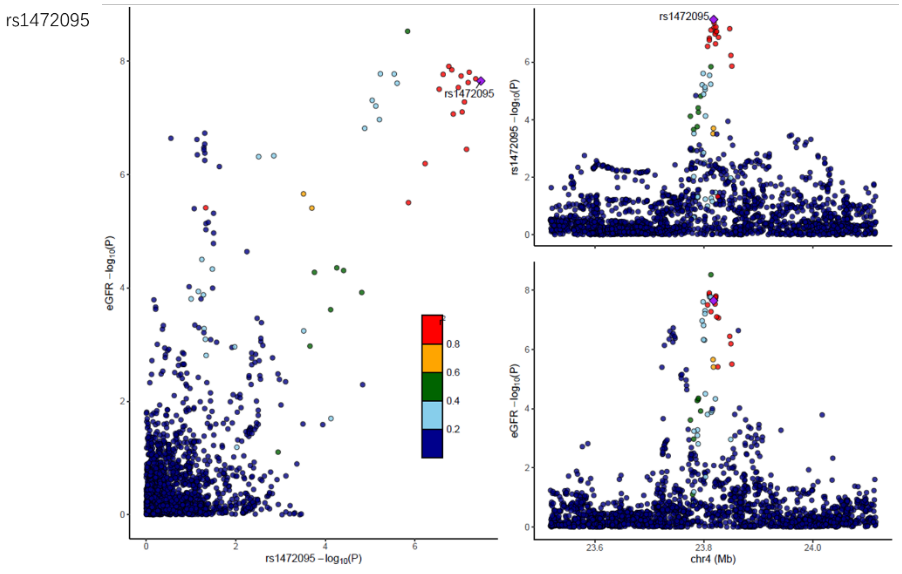


**Supplementary Figure 6**.

Plots showing high colocalization relastionship (i.e., PP.$H_4$ > 0.8) between rs1472095 and estimated glomerular filtration rate (pmid: 31152163).

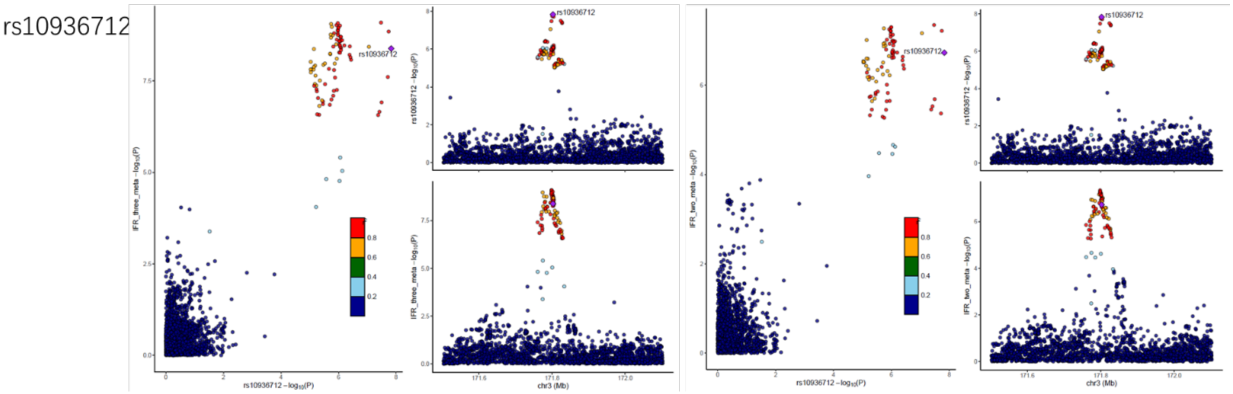


**Supplementary Figure 7**.

Plots showing high colocalization relastionship between rs10936712, immature fraction of reticulocytes (three-way meta) and immature fraction of reticulocytes (two-way meta) (pmid: 27863252)

rs13031826

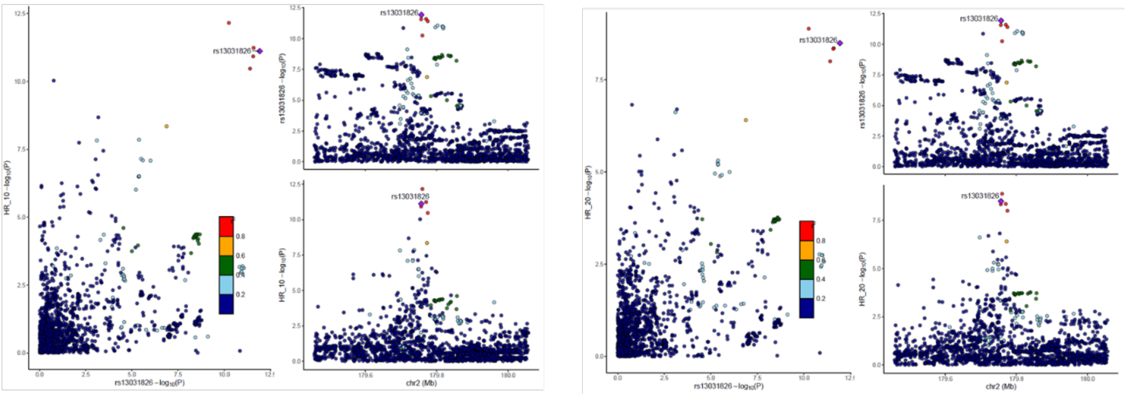

**Supplementary Figure 8**.

Plots showing high colocalization relastionship between rs13031826 and heart rate recovery at 10 secnds and heart rate recovery at 20 secnds (pmid: 29497042)

rs7433306

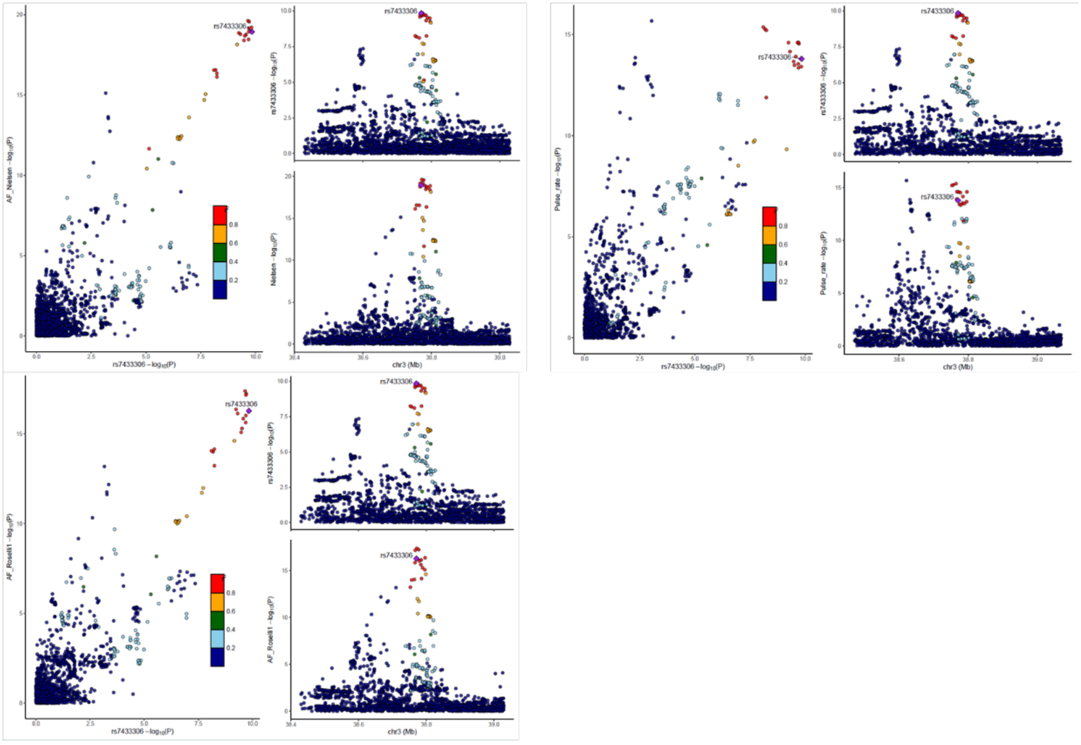

**Supplementary Figure 9**.

Plots showing high colocalization relastionship between rs7433306, atrial fibrillation (pmid: 29497042 and 29892015), and pulse rate (pmid: 31427789).

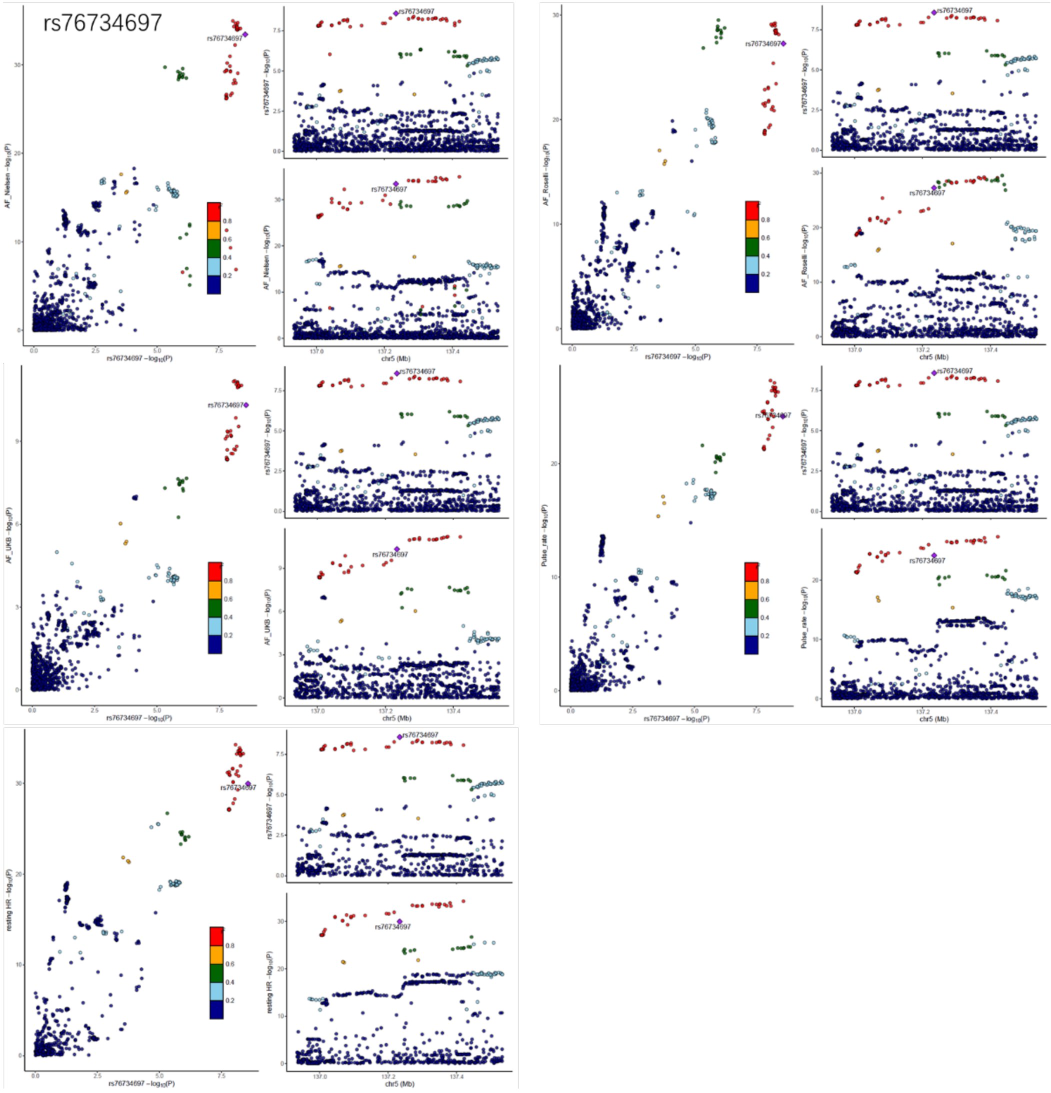


**Supplementary Figure 10**.

Plots showing high colocalization relastionship between rs76734697, atrial fibrillation (pmid: 29497042, 31427789 and 29892015), and resting heart rate (pmid: 30940143) and pulse rate (pmid: 31427789).

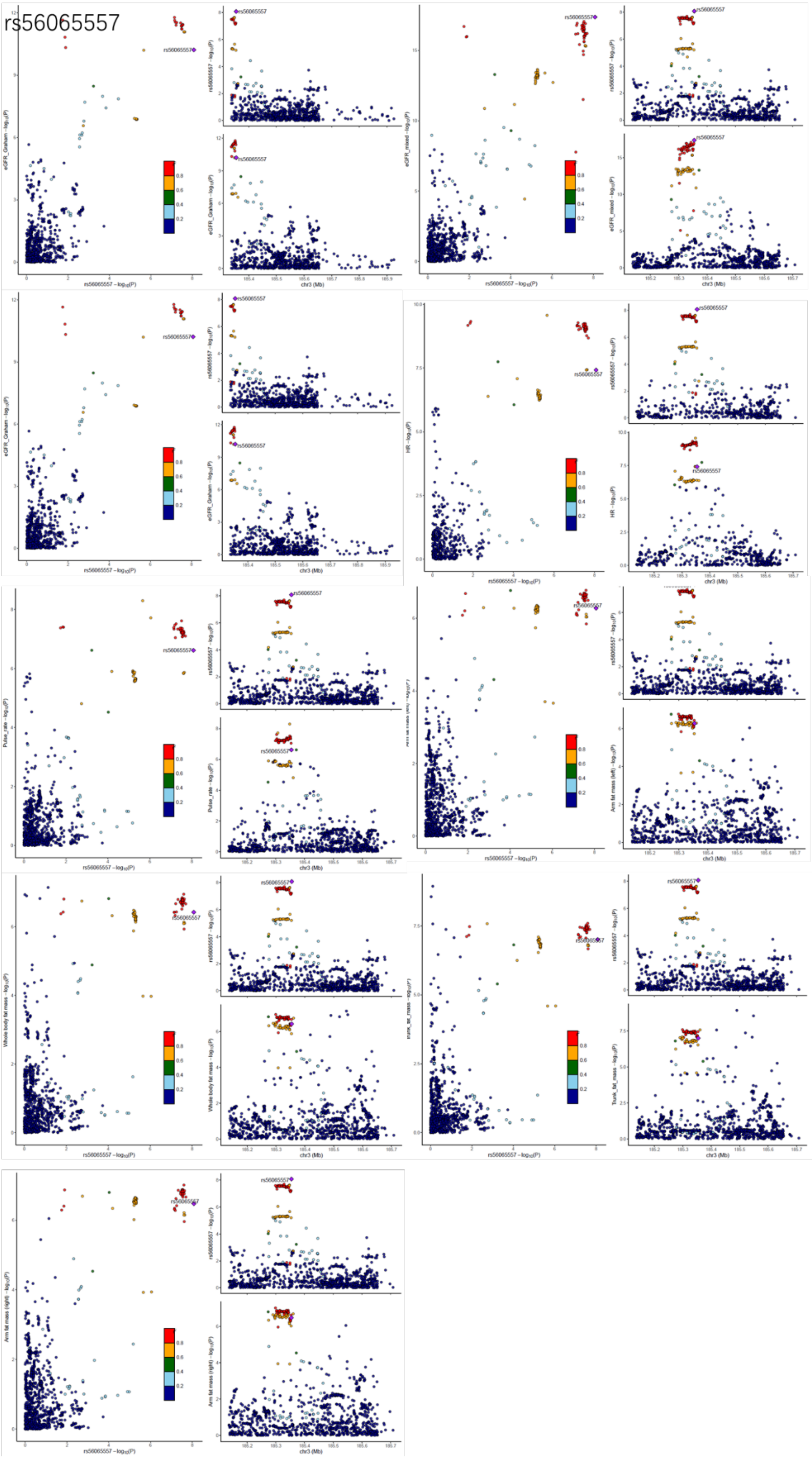


**Supplementary Figure 11**.

Plots showing high colocalization relastionship between rs56065557, Estimated glomerular filtration rate (pmid: 31152163, both European and mixed ancestery; 31015462), resting heart rate (pmid: 30940143), pulse rate (pmid: 31427789), trunk fat mass, right arm fat mass, left arm fat mass and whole body fat mass (pmid: 31427789).
